# COVID-19 impact on routine immunisations for vaccine-preventable diseases: Projecting the effect of different routes to recovery

**DOI:** 10.1101/2022.01.07.22268891

**Authors:** Jaspreet Toor, Xiang Li, Mark Jit, Caroline L Trotter, Susy Echeverria-Londono, Anna-Maria Hartner, Jeremy Roth, Allison Portnoy, Kaja Abbas, Neil M Ferguson, Katy AM Gaythorpe

## Abstract

**Introduction:** Over the past two decades, vaccination programmes for vaccine-preventable diseases (VPDs) have expanded across low- and middle-income countries (LMICs). However, the rise of COVID-19 resulted in global disruption to routine immunisation (RI) activities. Such disruptions could have a detrimental effect on public health, leading to more deaths from VPDs, particularly without mitigation efforts. Hence, as RIs resume, it is important to estimate the effectiveness of different approaches for recovery.

**Methods:** We apply an impact extrapolation method developed by the Vaccine Impact Modelling Consortium to estimate the impact of COVID-19-related disruptions with different recovery scenarios for ten VPDs across 112 LMICs. We focus on deaths averted due to RIs occurring in the years 2020-2030 and investigate two recovery scenarios relative to a no-COVID-19 scenario. In the recovery scenarios, we assume a 10% COVID-19-related drop in RI coverage in the year 2020. We then linearly interpolate coverage to the year 2030 to investigate two routes to recovery, whereby the immunization agenda (IA2030) targets are reached by 2030 or fall short by 10%.

**Results:** We estimate that falling short of the IA2030 targets by 10% leads to 11.26% fewer fully vaccinated persons (FVPs) and 11.34% more deaths over the years 2020-2030 relative to the no-COVID-19 scenario, whereas, reaching the IA2030 targets reduces these proportions to 5% fewer FVPs and 5.22% more deaths. The impact of the disruption varies across the VPDs with diseases where coverage expands drastically in future years facing a smaller detrimental effect.

**Conclusion:** Overall, our results show that drops in RI coverage could result in more deaths due to VPDs. As the impact of COVID-19-related disruptions is dependent on the vaccination coverage that is achieved over the coming years, the continued efforts of building up coverage and addressing gaps in immunity are vital in the road to recovery.

**SUMMARY:** *What is already known?:*

- The impact of vaccination programmes without COVID-19-related disruption has been assessed by the Vaccine Impact Modelling Consortium.
- The COVID-19 pandemic has disrupted vaccination programmes resulting in a decline in coverage in the year 2020, the ramifications of this is unclear.

*What are the new findings?:*

- We estimate the impact of disruptions to routine immunisation coverage and different routes to recovery. We compare to a scenario without COVID-19-related disruptions (assuming no drops in immunisation coverage).
- We estimate that reaching the Immunization Agenda (IA2030) targets leads to 5% fewer FVPs and 5.22% more deaths over the years 2020 to 2030 relative to the scenario with no COVID-19-related disruptions, whereas falling short of the IA2030 targets by 10% leads to 11.26% fewer fully vaccinated persons (FVPs) and 11.34% more deaths.
- The impact of the disruption varies across the vaccine-preventable diseases with those forecasted to have vast expansions in coverage post-2020 able to recover more.

*What do the new findings imply?:*

- A drop in vaccination coverage results in fewer vaccinated individuals and thus more deaths due to vaccine-preventable diseases. To mitigate this, building up coverage of routine immunisations and addressing immunity gaps with activities such as catch-up campaigns are vital in the road to recovery.

## 1 INTRODUCTION

The coronavirus disease-19 (COVID-19) pandemic has resulted in disruption to health services globally, including disruption of vaccination activities. Many low- and middle-income countries (LMICs) have faced drops in routine immunisation (RI) coverage, alongside delays in supplementary immunisation activities (SIAs, such as campaigns).^1,2^

In response to the evolving pandemic, early World Health Organization (WHO) guidance in March 2020 recommended temporary suspension of mass vaccination campaigns but continuation of RIs whilst maintaining prevention and control measures for COVID-19.^3^ Later, in May 2020, WHO interim guidance recommended consideration of vaccine-preventable disease (VPD) outbreak risk when deciding whether to conduct campaigns.^4^ The WHO’s latest national pulse survey on the continuity of essential health services during the COVID-19 pandemic highlights that there has been a reduction in the number of countries (37% of 112 responding countries) reporting disruptions to immunisation services in 2021, compared to 62% of 129 countries reporting disruptions in 2020.^2^

The level of decline in vaccination coverage has been estimated throughout the pandemic. Early projections by the Institute for Health Metrics and Evaluation (IHME) estimated the disruption that had already occurred until July 2020 (based on various data sources, including survey data, monthly administrative data on health services and data on human mobility patterns) and projected what may occur for the remainder of 2020.^5^ This led to a prediction of a 7-17% drop in coverage of third diphtheria-tetanus-pertussis (DTP3) dose in 2020.^6^ Further modelling work by IHME used administrative data and reports from electronic immunisation systems, with mobility data as a model input and estimated the global coverage in 2020 for DTP3 and the first dose of measles-containing vaccine (MCV1) to have fallen by 7.7% and 7.9%, respectively.^7^ Notably, the level of disruption varies geographically with some areas more affected than others.^7,8^ In Gavi-supported countries, coverage with a full course of pentavalent vaccine and MCV1 both decreased to 78% from 82% and 81%, respectively.^8^ Despite disruptions, more children were vaccinated in December 2020 than in December 2019, showcasing the vast efforts made by countries and international organisations to bring vaccination programmes back on track.^8^

Prior to the pandemic, vaccination coverage for VPDs had been increasing, significantly reducing morbidity and mortality related to VPDs across LMICs.^9^ In LMICs, the immense progress made has been in part due to Gavi, the Vaccine Alliance, which was created in 2000 with a goal of providing vaccines to save lives and protect people’s health. Over 2000-2020, Gavi has supported vaccination for over 888 million children through routine programmes and over 1.19 billion vaccinations through preventive vaccination campaigns, preventing over 15 million future deaths.^10^

The beneficial population-level effects of vaccination programmes cannot be assessed directly as the counterfactual, i.e. the situation without vaccination, cannot be observed. Advantageously, mathematical models enable us to quantify the impact of vaccination in terms of cases, deaths and disability-adjusted life years averted. The Vaccine Impact Modelling Consortium (VIMC), established in 2016, consists of multiple independent modelling groups with the aim of estimating the impact of vaccination programmes for 12 VPDs over 112 LMICs.^11^ Recently, the VIMC estimated that without COVID-19-related disruptions to vaccination coverage, 47 (95%CrI[39, 56]) million deaths would be averted due to vaccination activities over 2020–2030 for 10 VPDs across 112 LMICs.^9^

Furthermore, a study conducted to inform the Immunization Agenda (IA2030) estimated that 51.0 million (95% CI[48.5, 53.7]) deaths would be averted due to vaccinations administered between the years 2021 and 2030 for 14 pathogens in 194 countries.^12^ IA2030 is based on aspirational country-specific DTP3 2030 targets. The study assumed 2019 coverage levels remained constant through 2021.

A previous study assessing the impact of COVID-19 disruptions on VPDs, estimated the health impact of 50% reduced RI coverage in 2020 and delay of campaign vaccination from 2020 to 2021 for measles, MenA and YF showing risks of increased disease burden and measles outbreaks.^13^

It is important to estimate the long-term impact of different routes to recovery to determine their effectiveness. In this study, we investigate the impact of COVID-19-related disruptions with different recovery scenarios for ten VPDs, namely, *Haemophilus influenzae* type b (Hib), hepatitis B (HepB), human papillomavirus (HPV), Japanese encephalitis (JE), measles, *Neisseria meningitidis* serogroup A (MenA), rotavirus (Rota), rubella, *Streptococcus pneumoniae* (PCV) and yellow fever (YF). We use an impact extrapolation (IE) method developed by the VIMC to estimate the effect of RI coverage changes. Impact is attributed to the year in which the RI activity occurred.^14^ We examine a 10% drop in coverage in the year 2020 and linearly project different routes to recovery over the years 2021 to 2030 i.e. reaching IA2030 targets in the year 2030 or falling 10% short. Comparing this to a scenario with no COVID-19-related disruptions, we determine the effectiveness of the different routes to recovery.

## 2 METHODS

To estimate the effect of coverage changes to RIs, we use the IE method developed by the VIMC (Fig 1).^14^ This method is computationally and time efficient as it allows us to take the impact ratio (impact attributable per fully vaccinated person) from previous VIMC work and apply it to a new coverage scenario in order to extrapolate the impact calculation to the new coverage estimates. More specifically, the new number of fully vaccinated persons (FVPs) is first calculated by multiplying the new coverage with the target population size. Then to calculate the updated impact of vaccination, we multiply previously estimated impact ratios associated with RIs^9^ with the new number of FVPs (Fig 1).

**Figure 1:**
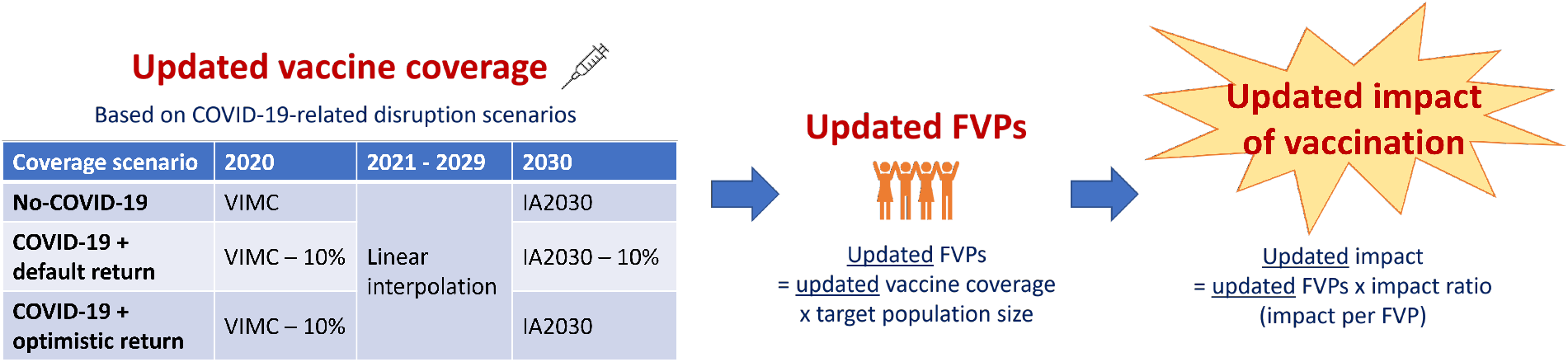
Schematic of impact extrapolation (IE) method and coverage scenarios over the years 2020 to 2030. Vaccine Impact Modelling Consortium (VIMC) coverage corresponds to coverage used for the previous VIMC work^9^ and IA2030 coverage corresponds to the immunization agenda coverage.^12^

As the IE method is primarily a linear interpolation of previous vaccine impact estimates with new coverage estimates, it may not capture any outbreaks or existing population immunity that may occur for the outbreak-prone VPDs with dynamic models (Table 1).^14^

**Table 1:** For the ten vaccine-preventable diseases analysed: number of countries with routine immunisation activities over the years 2020 to 2030; type of model(s) used by the Vaccine Impact Modelling Consortium; type of vaccination activities (routine immunisation, RI, and/or supplementary immunisation activities, SIA); risk of outbreaks occurring.

| Disease | Number of countries | Model type(s) | Activity type(s) | Outbreak risk |
| --- | --- | --- | --- | --- |
| *Haemophilus influenzae* type b (Hib) | 2020-2021: 110, 2022-2030: 112 | 2 static | RI | Minimal |
| Hepatitis B (HepB) | 2020-2030: 112 | 2 dynamic + 1 static | RI | Some |
| Human papillomavirus (HPV) | 2020-2022: 17, 2023-2030: 110 | 2 static | RI + SIA | Minimal |
| Japanese encephalitis (JE) | 2020-2022: 8, 2023-2030: 17 | 1 dynamic + 1 static | RI + SIA | Minimal |
| Measles | 2020-2030: 112 | 2 dynamic | RI + SIA | Yes |
| *Neisseria meningitidis* serogroup A (MenA) | 2020-2030: 26 | 2 dynamic | RI + SIA | Yes |
| Rotavirus (Rota) | 2020-2021: 64, 2022-2030: 112 | 1 dynamic + 1 static | RI | Yes |
| Rubella | 2020-2022: 88, 2023-2030: 112 | 2 dynamic | RI + SIA | Yes |
| *Streptococcus pneumoniae* (PCV) | 2020-2030: 112 | 2 static | RI | Minimal |
| Yellow fever (YF) | 2020-2023: 25, 2024-2030: 36 | 2 static | RI + SIA | Yes |

Impact is attributed to the year in which the vaccination activity took place (year of vaccination activity stratification method^14^). This method estimates the long-term impact of vaccination due to RIs that occur in a particular year. Hence, we capture the impact for pathogens where the mortality occurs later in life, such as HepB and HPV. Here, we focus on impact in terms of deaths averted due to RIs that occur over the years 2020 to 2030. Using the IE with the year of vaccination activity stratification method, the effect of coverage improvements is averaged over the whole time period.^14^

In the previous VIMC model runs, twenty-one mathematical models were used to inform the estimates with two models per pathogen (except HepB which has three models) thereby increasing robustness as the models differ in their underlying assumptions and modelling frameworks. The previous estimates did not include COVID-19-related disruptions to coverage. Model estimates focused on 112 countries, including 73 currently and formerly Gavi supported countries and 39 other countries that are of interest due to high burden and/or potential vaccine introduction.^9^ Pathogens endemic only in certain regions such as JE, MenA, and YF have estimates for 17, 26 and 36 countries, respectively (Table 1). Standardised, national-level, age-stratified demographic data were provided to all modellers from the 2019 United Nations World Population Prospects (UNWPP) for years 2000 to 2100.^15^ Detailed model descriptions are provided in^9^ (HepB,^16^ HPV,^17,18^ Hib,^19,20^ JE,^21^ Measles,^22,23^ MenA,^24,25^ PCV,^19,20^ Rota,^26,27^ Rubella,^28,29^ YF^30^).

We investigate three scenarios: one with no COVID-19-related disruptions and two with COVID-19-related disruptions (Fig 1). Coverage estimates for 2020 are based on the previous VIMC model estimates for all 112 countries.^9^ For the no-COVID-19 scenario, we assume no drop in RI coverage in the year 2020 and linearly interpolate coverage over 2021 to 2029, assuming that the IA2030 targets are reached in the year 2030. For the disruption scenarios, we assume a 10% drop in RI coverage in the year 2020 then linearly interpolate coverage over 2021 to 2029, assuming that coverage in 2030 either reaches the IA2030 targets (IA2030 return scenario) or reaches the IA2030 targets with a 10% absolute reduction (default return scenario).^12^

Across all scenarios, we assume vaccine introduction years remain the same. Where RIs were introduced later than 2020, we keep the coverage the same as it was expected to be in the year of introduction and linearly interpolate to the scenario-specific 2030 endpoint.

To compare the disruption scenarios to the no-COVID-19 scenario, we estimate the change in FVPs and deaths averted for each disease. More specifically, we show the proportional decrease in FVPs and increase in deaths in the COVID-19 scenarios relative to the no-COVID-19 scenario. We also estimate the impact (deaths averted) and FVPs attributable to each years RI activities over the years 2020-2030. Note that the death terms in the COVID-19 scenarios do not include deaths due to COVID-19, only deaths attributable to the ten VPDs analysed.

### 2.1 Patient and Public Involvement

Patients and the public were not involved in this study.

## 3 RESULTS

We compare the two recovery scenarios to the scenario without COVID-19-related disruptions, focusing on the relative change in FVPs and deaths averted due to RIs occurring over the years 2020 to 2030 for the ten diseases. With the default return scenario over 2020-2030, we estimate that a COVID-19 drop in coverage leads to 11.26% fewer FVPs and 11.34% more deaths relative to the no-COVID-19 scenario. With the IA2030 return scenario over 2020-2030, these proportions decline to 5% fewer FVPs and 5.22% more deaths (Table 2).

**Table 2:** Fully vaccinated persons (FVPs) and deaths averted due to routine immunisation activities over the years 2020-2030 for each disease and in total across all ten diseases (*Haemophilus influenzae* type b (Hib), hepatitis B (HepB), human papillomavirus (HPV), Japanese encephalitis (JE), measles, *Neisseria meningitidis* serogroup A (MenA), rotavirus (Rota), rubella, *Streptococcus pneumoniae* (PCV) and yellow fever (YF)). Relative change (%) in FVPs and deaths averted in comparison to the no-COVID-19 scenario also shown where relative change is given by 100*(COVID-19 scenario - no-COVID-19 scenario)/no-COVID-19 scenario. See Fig 1 for more detail on the coverage scenarios.

| Coverage scenario | Disease | FVPs in millions<br>(Relative change (%)) | Deaths averted in millions<br>(Relative change (%)) |
| --- | --- | --- | --- |
| **No-COVID-19** | HepB | 1890.67 | 12.55 |
| 2020: VIMC | Hib | 1037.56 | 2.45 |
| 2021-2029: Linear interpolation | HPV | 322.28 | 3.84 |
| 2030: IA2030 | JE | 261.37 | 0.12 |
|  | Measles | 2111.43 | 19.32 |
|  | MenA | 225.48 | 0.19 |
|  | PCV | 917.14 | 2.10 |
|  | Rota | 853.42 | 0.66 |
|  | Rubella | 2014.30 | 0.65 |
|  | YF | 243.99 | 1.84 |
|  | Total | 9877.65 | 43.72 |
| **COVID-19 + default return** | HepB | 1683.16 (-10.98) | 11.14 (-11.21) |
| 2020: VIMC -10% | Hib | 923.25 (-11.02) | 2.16 (-11.73) |
| 2021-2029: Linear interpolation | HPV | 299.37 (-7.11) | 3.55 (-7.50) |
| 2030: IA2030 - 10% | JE | 216.20 (-17.28) | 0.10 (-15.61) |
|  | Measles | 1876.60 (-11.12) | 17.03 (-11.86) |
|  | MenA | 190.44 (-15.54) | 0.16 (-15.03) |
|  | PCV | 792.56 (-13.58) | 1.83 (-13.16) |
|  | Rota | 759.96 (-10.95) | 0.59 (-10.98) |
|  | Rubella | 1811.87 (-10.05) | 0.59 (-9.93) |
|  | YF | 211.70 (-13.23) | 1.61 (-12.33) |
|  | Total | 8765.12 (-11.26) | 38.76 (-11.34) |
| **COVID-19 + IA2030 return** | HepB | 1795.86 (-5.02) | 11.86 (-5.49) |
| 2020: VIMC -10% | Hib | 984.18 (-5.14) | 2.31 (-5.66) |
| 2021-2029: Linear interpolation | HPV | 321.13 (-0.36) | 3.82 (-0.70) |
| 2030: IA2030 | JE | 243.78 (-6.73) | 0.12 (-5.99) |
|  | Measles | 1999.97 (-5.28) | 18.21 (-5.73) |
|  | MenA | 208.50 (-7.53) | 0.17 (-7.30) |
|  | PCV | 855.04 (-6.77) | 1.97 (-6.48) |
|  | Rota | 817.68 (-4.19) | 0.64 (-3.93) |
|  | Rubella | 1926.59 (-4.35) | 0.63 (-4.25) |
|  | YF | 230.46 (-5.55) | 1.73 (-5.82) |
|  | Total | 9383.19 (-5) | 41.44 (-5.22) |

The impact of the COVID-19-related disruption varies across the diseases with some diseases able to recover more FVPs and deaths averted. With both recovery options, HPV is estimated to face the lowest effect due to disruption relative to the other VPDs. More specifically, HPV has a 0.36% or 7.11% reduction in FVPs and 0.7% or 7.5% more deaths with the IA2030 or default return, respectively (Table 2 and Fig 2). HPV RIs are introduced into many countries in 2023 (Table 1), prior to that RIs are only present in 17 countries. RIs occurring in 2020 contribute to less than 1% of the FVPs and deaths averted over the years 2020 to 2030 (Fig 3). Hence, as a larger proportion of impact due to HPV RIs occurs post-2022, the COVID-19-related decline in coverage in the year 2020 has a relatively low impact. RIs in the year 2030 contribute to the largest proportion of impact, 14.91% of deaths averted, for HPV (Fig 3). With the IE and the year of vaccination activity stratification method, the later improvements in coverage are being averaged out over the whole time period. Hence, the beneficial effects of earlier HPV RIs may be artificially inflated, thereby contributing to the low estimate of disruption impact.

**Figure 2:**
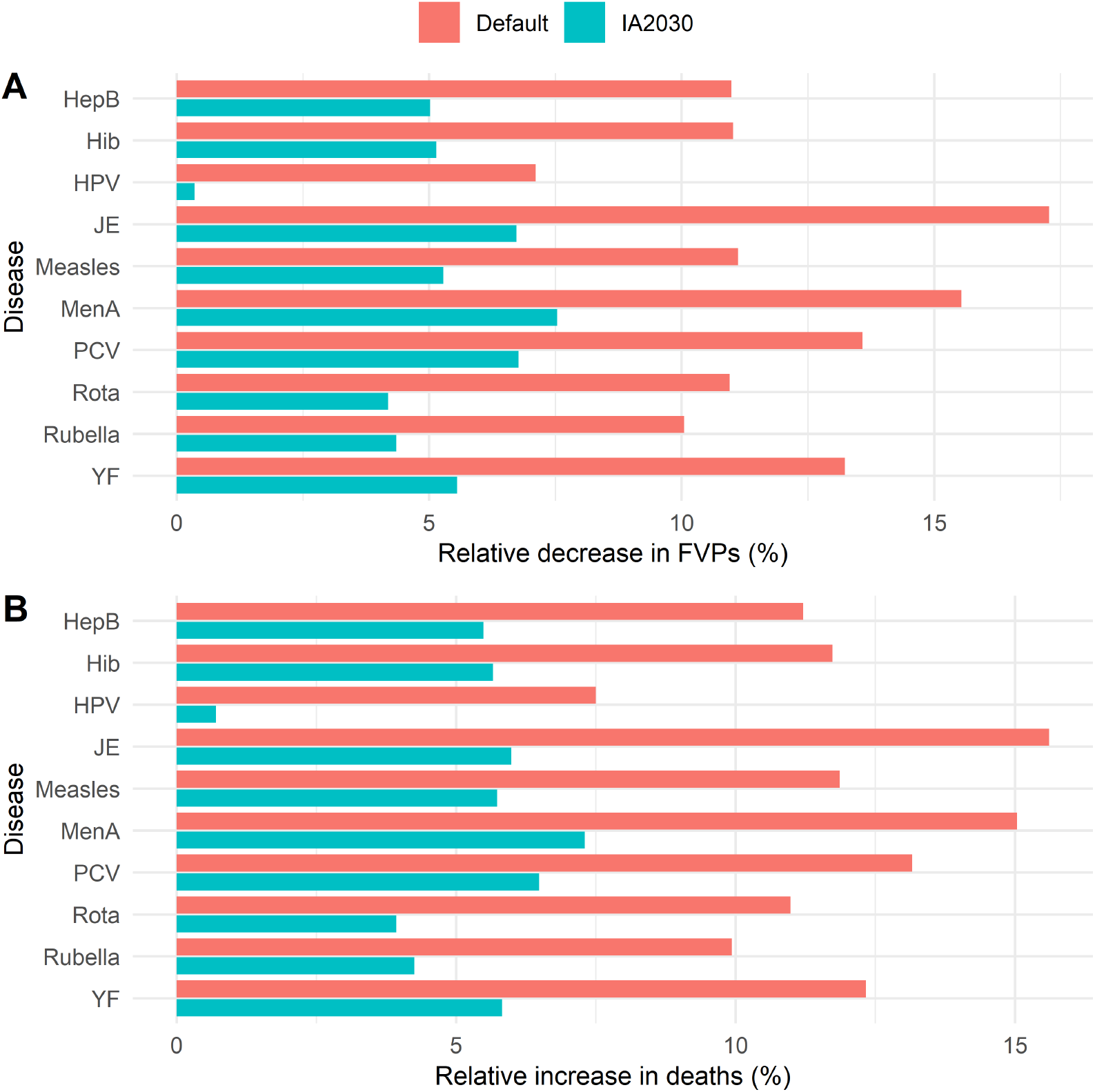
Change over the years 2020-2030 for each disease in (A) fully vaccinated persons (FVPs) and (B) deaths in the default and IA2030 scenarios relative to the no-COVID-19 scenario. Disease abbreviations: *Haemophilus influenzae* type b (Hib), hepatitis B (HepB), human papillomavirus (HPV), Japanese encephalitis (JE), measles, *Neisseria meningitidis* serogroup A (MenA), rotavirus (Rota), rubella, *Streptococcus pneumoniae* (PCV) and yellow fever (YF). See Table 2 for more detailed information.

**Figure 3:**
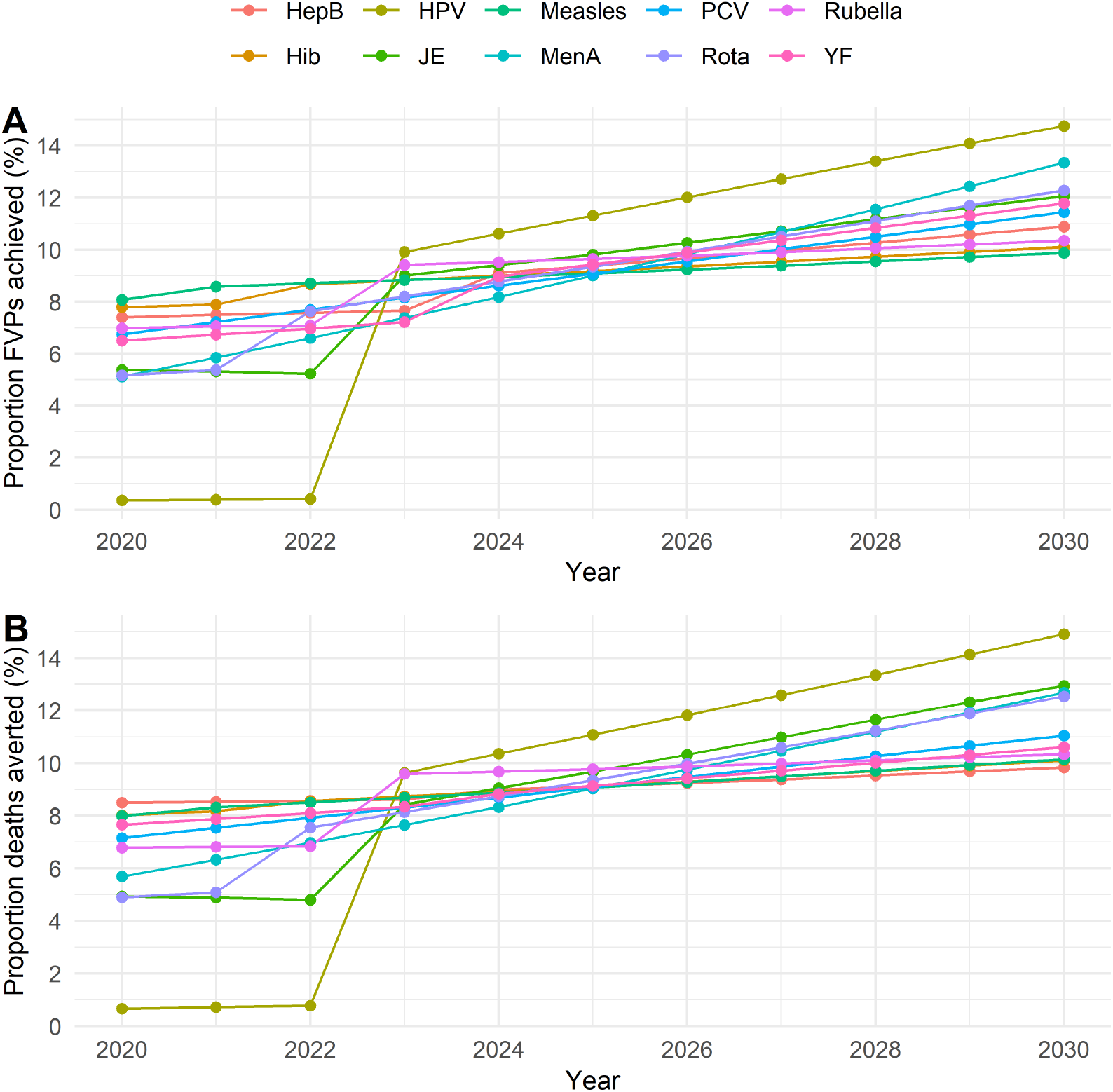
Proportional (A) fully vaccinated persons (FVPs) and (B) impact (in terms of deaths averted) attributable to each year’s RI activities in the no-COVID-19 scenario for *Haemophilus influenzae* type b (Hib), hepatitis B (HepB), human papillomavirus (HPV), Japanese encephalitis (JE), measles, *Neisseria meningitidis* serogroup A (MenA), rotavirus (Rota), rubella, *Streptococcus pneumoniae* (PCV) and yellow fever (YF).

On the other hand, we estimate that JE and MenA face the largest relative effects due to disruption. Similar to HPV, JE only has RIs in 8 countries in 2020. However, JE RIs remain in relatively fewer (17) countries by 2030. MenA RIs are also present in relatively fewer (26) countries over the years 2020 to 2030. The impact attributable to each year of RIs is similar for these VPDs as it increases from 4.94-5.69% of deaths averted attributable to RIs in 2020 to 12.68-12.95% of deaths averted attributable to RIs in 2030 (Fig 3). With the default return, MenA and JE have the largest impacts due to disruption with a 15.54-17.28% drop in FVPs and 15.03-15.61% more deaths, respectively (Table 2). As JE and MenA RIs in the year 2030 contribute to a large proportion of the deaths averted over 2020-2030, the shortfall of 10% in the default return scenario has a large impact. Moving to the IA2030 return, the drop in FVPs declines to 6.73-7.53% with 5.99-7.30% more deaths for JE and MenA, respectively (Table 2 and Fig 2).

Relative to the other VPDs, PCV and YF face a moderate impact due to COVID-19-related disruptions. They have a similar pattern of the proportion of FVPs achieved and death averted attributable to each year of RIs with the proportion of deaths averted increasing stably from 7.16-7.66% attributable to 2020 RIs to 10.61-11.05% attributable to 2030 RIs (Fig 3). With the default return, these VPDs range between a 13.23-13.58% drop in FVPs and 12.33-13.16% increase in deaths relative to the no-COVID-19 scenario. With the IA2030 return, these ranges decline to a 5.55-6.77% drop in FVPs and 5.82-6.48% increase in deaths (Table 2 and Fig 2). Note that this does not capture any YF outbreaks.

For the VPDs that have RIs over a large number of countries from the years 2020 to 2030, i.e. HepB, Hib and measles, the impact of the COVID-19-related disruption is similar (Table 1). For these VPDs, the FVPs achieved and deaths averted attributable to each year’s RIs over 2020-2030 is more evenly distributed (with proportion of deaths averted ranging from 7.99-8.5% attributable to 2020 RIs to 9.83-10.15% attributable to 2030 RIs; Fig 3). With the default return, these VPDs range between a 10.98-11.17% drop in FVPs and 11.21-11.86% increase in deaths relative to the no-COVID-19 scenario. With the IA2030 return, these ranges decline to a 5.02-5.28% drop in FVPs and 5.49-5.73% increase in deaths (Table 2 and Fig 2).

Rota and rubella also have coverage in all 112 countries by 2030, highlighted by the increase in proportional FVPs and impact attributable to each year of RIs (4.89-6.78% deaths averted in 2020 and 10.34-12.53% deaths averted in 2030; Table 1 and Fig 3). With the default return, these diseases range between a 10.05-10.95% drop in FVPs and 9.93-10.98% increase in deaths relative to the no-COVID-19 scenario. With the IA2030 return, these ranges decline to a 4.19-4.35% drop in FVPs and 3.93-4.25% increase in deaths (Table 2 and Fig 2).

It is important to note that although we focus on proportional decrease in FVPs and increase in deaths for each of the VPDs analysed, a specific proportional change is not equivalent across the VPDs as the baseline numbers are different. For example, a 1% increase in deaths attributable to measles corresponds to a larger number of deaths than a 1% increase in deaths attributable to a VPD across fewer countries, such as JE (baseline numbers shown in Table 2).

Overall, the IA2030 return scenario reduces the impact of COVID-19-related disruption for each of the VPDs. The impact of the disruption varies by VPD, with HPV facing the lowest impact as the year 2020 is attributable to a relatively low proportion of deaths averted over the years 2020 to 2030. This is dependent on drastic expansions in coverage which faces a risk of vaccine introduction delays or cancellations. VPDs, such as JE and MenA, where the year 2030 is attributable to a relatively high proportion of deaths averted over the years 2020 to 2030 are at risk if high coverage is not achieved by 2030. Furthermore, the outbreak-prone VPDs may be at higher risk than estimated though any existing immunity from earlier campaigns may aid in reducing risks.

### 3.1 Outbreak-prone VPDs: Comparison to previous VIMC work

As the IE method risks not capturing any outbreaks for some of the VPDs (Table 1), we compare the results of our IE analysis to modelling work carried out by VIMC members for measles, MenA and YF.^13^ Note: both VIMC models for YF are static so even with new model estimates, YF outbreaks may not be captured. The model estimates for measles showed that the risk of an outbreak due to delayed campaigns varied by country, with those that had high pre-existing immunity facing lower risks of outbreaks. For MenA, in areas where there have been recent introductory campaigns capturing a wide range of ages (ages 1 to 29), there is more persistence of direct and indirect benefits which mitigate the impact of disruptions. Similarly for YF, strong existing herd immunity in areas where there have been recent campaigns aids in reducing the impact of disruptions.^13^

Our IE analysis does not take into account the impact of previous campaigns so any existing immunity for VPDs such as measles, MenA and YF is not being captured. Hence, our IE analysis may be overestimating the impact of COVID-19-related disruptions for areas where there is immunity from previous vaccination activities. This is the case particularly for MenA as, in contrast to the model estimates, the IE estimated it to be one of the most impacted VPDs. For outbreak-prone VPDs that are modelled dynamically, the IE results should be treated carefully with campaigns and immunity in mind. New model runs (though more computationally expensive) would be required to account for these factors.

## 4 DISCUSSION

In this study, we have used the IE method, extrapolating previous VIMC impact ratios to new coverage scenarios based on COVID-19-related disruptions. We have found that declines in RI coverage result in fewer FVPs and more deaths. This can be mitigated by achieving high coverage levels, with achievement of the IA2030 targets resulting in 5% fewer FVPs and 5.22% more deaths than the no-COVID-19 scenario.

We have also found that the impact of the disruption varies by VPD with HPV estimated to face a relatively lower impact than the other VPDs, driven by the large expansions in coverage in future years (0.7% increase in deaths with the IA2030 return). However, the impact of the COVID-19 pandemic on HPV may be higher with delays or cancellations of HPV vaccine introductions and school closures. The other VPDs face a moderate to high impact, with JE and MenA estimated to have relatively higher impacts particularly if the IA2030 targets are not reached (15.03-15.61% more deaths in the default return reduced to 5.99-7.30% more deaths in the IA2030 return). In areas where there is existing immunity from earlier vaccination activities, these disruption impacts may be lower than estimated, particularly as our analysis is not accounting for any impact attributable to earlier campaigns that may have occurred.

Due to the IE methodology used, we have focused on proportional changes in FVPs and deaths for each of the VPDs analysed. However, a specific proportional change is not equivalent across the VPDs. As measles RIs are estimated to avert the largest number of deaths over the years 2020-2030, an increase in deaths attributable to measles due to declines in coverage corresponds to a larger number of deaths compared to the other VPDs.

We have implemented the IE method which is computationally and time efficient as it does not require new model estimates. The IE method has been shown to perform well for small changes to RI coverage but becomes less accurate when larger changes are made, including when SIAs are delayed or cancelled. Furthermore, any uncertainties in the previous model runs are propagated into the IE.^14^ Notably, for the outbreak-prone VPDs (measles, MenA, Rota, Rubella and YF), the IE method may be overestimating the impact of COVID-19-related disruptions for areas where there is existing immunity from earlier vaccination activities. Conversely, the IE method risks not capturing any outbreaks that may have occurred. With larger delays to activities, the risks of not capturing such outbreaks increases. To reduce inaccuracies in our analysis, we have limited coverage changes to small coverage changes to RI-only activities. In future work, to more accurately assess larger changes to coverage for both RIs and SIAs, and capture herd effects and possible outbreaks, new model estimates would be required.

Six of the VPDs assessed in our study (HPV, JE, measles, MenA, rubella and YF) have SIAs in addition to RIs which have not been incorporated here. A previous study showed that delaying campaigns may increase the risk of measles outbreaks and the disease burden for YF.^13^ With declines in vaccination coverage and in outbreak detection and control (which were reported as disrupted by 41% of 123 countries in 2020^2^), the risk of outbreaks increased for the outbreak-prone VPDs. By June 2020, Gavi reported that 30 Gavi eligible countries had reported VPD outbreaks, particularly for measles.^31^ In August 2020, the death toll from a measles outbreak in the Democratic Republic of the Congo surpassed 7,000 (the world’s worst).^32^

Importantly, a study promoting continuation of campaigns during the pandemic showed that personal protective equipment and symptomatic screening can reduce COVID-19 transmission.^33^ Hence, catch-up campaigns for these VPDs where delays or cancellations have occurred may aid in reducing immunity gaps and limiting the impact of COVID-19-related disruptions.^34^ In the latest WHO pulse survey, catch-up campaigns were reported by 31% of 112 countries as a mitigation strategy.^2^

In our scenarios, we have assumed the same level of disruption to coverage for all countries and VPDs examined. However, levels of disruption are likely to vary across countries and diseases. For example, the most severe reductions in DTP3 and MCV1 coverage in 2020 were seen in north Africa and the Middle East, south Asia, and Latin America and the Caribbean, with the lowest reductions in sub-Saharan Africa.^7^

With the global disruptions to vaccination programmes, there is evidence that the population of zero-dose children, i.e. children who have not received any dose of DTP, has increased. WHO and United Nations Children’s Fund (UNICEF) estimates indicate that the number of children who did not complete the 3-dose DTP series worldwide increased by 20% to 22.7 million from 2019 to 2020. Of these children, 17.1 million (75%) are estimated to be zero-dose children.^35^ To address such immunity gaps caused by disruptions, catch-ups and monitoring are important, e.g. through catch-up campaigns, expanding to reach age groups that may have aged out of the targeted ages and screening children in schools/health facilities for vaccination status.^35^

Furthermore, the COVID-19 pandemic has had widespread impacts on other aspects of society, such as school closures. In May 2020, the World Bank highlighted that schools had closed in 180 countries, with 85% of students worldwide out of school.^36^ Due to this, vaccines, such as HPV, which are heavily reliant on delivery in schools may be more negatively effected than assumed in our analysis. Conversely, there is also evidence that the rise in non-pharmaceutical interventions (e.g. social distancing) may reduce the transmission of certain pathogens, such as those that cause bacterial meningitis.^37^ Further work is required to assess the full effects of these interventions.

As there has been immense progress to bring vaccination programmes back on track,^8^ we have only considered disruption in the year 2020 but this may be prolonged in some countries. It is still difficult to envisage the level of disruption in 2021 onward that countries may face due to the ongoing pandemic. Despite this uncertainty, it remains clear that building up coverage and addressing immunity gaps will be vital over the coming years.

## 5 CONCLUSION

Our study shows the importance of building up coverage as vaccination programmes recover from disruptions caused by the COVID-19 pandemic. This will enable the achievement of more vaccinated individuals and reduce morbidity and mortality caused by VPDs over the future years.

## Data Availability

Data has been previously published and made available online by the Vaccine Impact Modelling Consortium (VIMC).

https://montagu.vaccineimpact.org/2021/visualisation/

## Acknowledgements

We would like to thank Kim Woodruff for helpful comments and project management aspects.

## References

[1] World Health Organization: Pulse survey on continuity of essential health services during the COVID-19 pandemic: interim report, 27 August 2020 (2020). https://www.who.int/publications/i/item/WHO-2019-nCoV-EHS_continuity-survey-2020.1

[2] World Health Organization: Second round of the national pulse survey on continuity of essential health services during the COVID-19 pandemic: January - March 2021. Interim report. (2021). https://www.who.int/publications/i/item/WHO-2019-nCoV-EHS-continuity-survey-2021.1

[3] World Health Organization: Guiding principles for immunization activities during the COVID-19 pandemic: interim guidance, 26 March 2020. World Health Organization. https://apps.who.int/iris/handle/10665/331590 Accessed 2021-10-21

[4] World Health Organization: Framework for decision-making: implementation of mass vaccination campaigns in the context of COVID-19. https://www.who.int/publications/i/item/{WHO}-2019-{nCoV}-{Framework_Mass_Vaccination}-2020.1 Accessed 2021-10-21

[5] Bill and Melinda Gates Foundation: 2020 Goalkeepers report: COVID-19, a global perspective. https://www.gatesfoundation.org/goalkeepers/downloads/2020-report/report_a4_en.pdf

[6] Goalkeepers 2020 report explore the data: Vaccines. https://www.gatesfoundation.org/goalkeepers/report/2020-report/progress-indicators/vaccines/

[7] Causey, K., Fullman, N., Sorensen, R.J.D., Galles, N.C., Zheng, P., Aravkin, A., Danovaro-Holliday, M.C., Martinez-Piedra, R., Sodha, S.V., Velandia-González, M.P., et al.: Estimating global and regional disruptions to routine childhood vaccine coverage during the covid-19 pandemic in 2020: a modelling study. The Lancet 398(10299), 522–534 (2021). doi:https://doi.org/10.1016/S0140-6736(21)01337-4

[8] Gavi, the Vaccine Alliance: Routine immunisation worldwide holds firm despite the pandemic. https://www.gavi.org/vaccineswork/routine-immunisation-worldwide-holds-firm-despite-pandemic

[9] Toor, J., Echeverria-Londono, S., Li, X., Abbas, K., Carter, E.D., Clapham, H.E., Clark, A., de Villiers, M.J., Eilertson, K., Ferrari, M., Gamkrelidze, I., Hallett, T.B., Hinsley, W.R., Hogan, D., Huber, J.H., Jackson, M.L., Jean, K., Jit, M., Karachaliou, A., Klepac, P., Kraay, A., Lessler, J., Li, X., Lopman, B.A., Mengistu, T., Metcalf, C.J.E., Moore, S.M., Nayagam, S., Papadopoulos, T., Perkins, T.A., Portnoy, A., Razavi, H., Razavi-Shearer, D., Resch, S., Sanderson, C., Sweet, S., Tam, Y., Tanvir, H., Tran Minh, Q., Trotter, C.L., Truelove, S.A., Vynnycky, E., Walker, N., Winter, A., Woodruff, K., Ferguson, N.M., Gaythorpe, K.A.: Lives saved with vaccination for 10 pathogens across 112 countries in a pre-COVID-19 world. eLife 10 (2021). doi:https://doi.org/10.7554/eLife.676. Accessed 2021-10-21

[10] Gavi, the Vaccine Alliance: Annual progress report 2020. Gavi, the Vaccine Alliance. https://www.gavi.org/sites/default/files/programmes-impact/our-impact/apr/Gavi-Progress-Report-2020.pdf Accessed 2021-10-21

[11] Imperial College London: Vaccine Impact Modelling Consortium. https://www.vaccineimpact.org/ Accessed 2021-10-21

[12] Carter, A., Msemburi, W., Sim, S.Y., A.M. Gaythorpe K., Lindstrand, A., Hutubessy, R.C.W.: Modeling the impact of vaccination for the immunization agenda 2030: Deaths averted due to vaccination against 14 pathogens in 194 countries from 2021-2030. Electronic Journal (2021). doi:https://doi.org/10.2139/ssrn.3830781. Accessed 2021-10-21

[13] Gaythorpe, K.A., Abbas, K., Huber, J., Karachaliou, A., Thakkar, N., Woodruff, K., Li, X., Echeverria-Londono, S., on COVID-19 Impact on Vaccine Preventable Disease, V.W.G., Ferrari, M., Jackson, M.L., McCarthy, K., Perkins, T.A., Trotter, C., Jit, M.: Impact of COVID-19-related disruptions to measles, meningococcal a, and yellow fever vaccination in 10 countries. eLife 10 (2021). doi:https://doi.org/10.7554/eLife.670. Accessed 2021-10-21

[14] Echeverria-Londono, S., Li, X., Toor, J., de-Villiers, M., Nayagam, S., Hallett, T.B., Abbas, K., Jit, M., Klepac, P., Jean, K., Garske, T., Ferguson, N.M., Gaythorpe, K.A.M.: How can the public health impact of vaccination be estimated? medRxiv (2021). doi:https://doi.org/10.1101/2021.01.08.21249378. Accessed 2021-02-11

[15] Prospects, W.P.: Population division - United Nations (2019). https://population.un.org/wpp/

[16] Nayagam, S., Thursz, M., Sicuri, E., Conteh, L., Wiktor, S., Low-Beer, D., Hallett, T.B.: Requirements for global elimination of hepatitis b: a modelling study. The Lancet Infectious Diseases 16(12), 1399–1408 (2016). doi:https://doi.org/10.1016/S1473-3099(16)30204-3. Accessed 2020-10-06

[17] Goldie, S.J., O’Shea, M., Campos, N.G., Diaz, M., Sweet, S., Kim, S.-Y.: Health and economic outcomes of HPV 16,18 vaccination in 72 GAVI-eligible countries. Vaccine 26(32), 4080–4093 (2008). doi:https://doi.org/10.1016/j.vaccine.2008.04.053. Accessed 2020-11-24

[18] Abbas, K.M., van Zandvoort, K., Brisson, M., Jit, M.: Effects of updated demography, disability weights, and cervical cancer burden on estimates of human papillomavirus vaccination impact at the global, regional, and national levels: a prime modelling study. The Lancet. Global health 8(4), 536–544 (2020). doi:https://doi.org/10.1016/S2214-109X(20)30022-X

[19] Clark, A., Jauregui, B., Griffiths, U., Janusz, C.B., Bolaños-Sierra, B., Hajjeh, R., Andrus, J.K., Sanderson, C.: Trivac decision-support model for evaluating the cost-effectiveness of haemophilus influenzae type b, pneumococcal and rotavirus vaccination. Vaccine 31 Suppl 3, 19–29 (2013). doi:https://doi.org/10.1016/j.vaccine.2013.05.045

[20] Walker, N., Tam, Y., Friberg, I.K.: Overview of the lives saved tool (list). BMC Public Health 13 Suppl 3, 1 (2013). doi:https://doi.org/10.1186/1471-2458-13-S3-S1. Accessed 2020-11-24

[21] Quan, T.M., Thao, T.T.N., Duy, N.M., Nhat, T.M., Clapham, H.: Estimates of the global burden of japanese encephalitis and the impact of vaccination from 2000-2015. eLife 9 (2020). doi:https://doi.org/10.7554/eLife.510. Accessed 2020-10-06

[22] Chen, S., Fricks, J., Ferrari, M.J.: Tracking measles infection through non-linear state space models. Journal of the Royal Statistical Society: Series C (Applied Statistics) 61(1), 117–134 (2012). doi:https://doi.org/10.1111/j.1467-9876.2011.01001.x. Accessed 2020-11-24

[23] Verguet, S., Johri, M., Morris, S.K., Gauvreau, C.L., Jha, P., Jit, M.: Controlling measles using supplemental immunization activities: a mathematical model to inform optimal policy. Vaccine 33(10), 1291–1296 (2015)

[24] Karachaliou, A., Conlan, A.J.K., Preziosi, M.-P., Trotter, C.L.: Modeling long-term vaccination strategies with MenAfriVac in the african meningitis belt. Clinical Infectious Diseases 61 Suppl 5, 594–600 (2015). doi:https://doi.org/10.1093/cid/civ508. Accessed 2020-11-24

[25] Tartof, S., Cohn, A., Tarbangdo, F., Djingarey, M.H., Messonnier, N., Clark, T.A., Kambou, J.L., Novak, R., Diomandé, F.V.K., Medah, I., Jackson, M.L.: Identifying optimal vaccination strategies for serogroup a neisseria meningitidis conjugate vaccine in the african meningitis belt. Plos One 8(5), 63605 (2013). doi:https://doi.org/10.1371/journal.pone.0063605. Accessed 2020-11-24

[26] Pitzer, V.E., Atkins, K.E., de Blasio, B.F., Van Effelterre, T., Atchison, C.J., Harris, J.P., Shim, E., Galvani, A.P., Edmunds, W.J., Viboud, C., Patel, M.M., Grenfell, B.T., Parashar, U.D., Lopman, B.A.: Direct and indirect effects of rotavirus vaccination: comparing predictions from transmission dynamic models. Plos One 7(8), 42320 (2012). doi:https://doi.org/10.1371/journal.pone.0042320. Accessed 2020-11-24

[27] Clark, A., Tate, J., Parashar, U., Jit, M., Hasso-Agopsowicz, M., Henschke, N., Lopman, B., Van Zandvoort, K., Pecenka, C., Fine, P., Sanderson, C.: Mortality reduction benefits and intussusception risks of rotavirus vaccination in 135 low-income and middle-income countries: a modelling analysis of current and alternative schedules. The Lancet. Global health 7(11), 1541–1552 (2019). doi:https://doi.org/10.1016/S2214-109X(19)30412. Accessed 2020-11-24

[28] Boulianne, N., De Serres, G., Ratnam, S., Ward, B.J., Joly, J.R., Duval, B.: Measles, mumps, and rubella antibodies in children 5-6 years after immunization: effect of vaccine type and age at vaccination. Vaccine 13(16), 1611–1616 (1995). doi:https://doi.org/10.1016/0264-410x(95)00098-l. Accessed 2020-11-24

[29] Vynnycky, E., Papadopoulos, T., Angelis, K.: The impact of measles-rubella vaccination on the morbidity and mortality from congenital rubella syndrome in 92 countries. Human vaccines & immunotherapeutics 15(2), 309–316 (2019). doi:https://doi.org/10.1080/21645515.2018.1532257. Accessed 2020-10-06

[30] Gaythorpe, K.A.M., Hamlet, A.T.P., Jean, K., Ramos, D.G., Cibrelus, L., Garske, T., Ferguson, N.M.: The global burden of yellow fever. medRxiv (2020). doi:https://doi.org/10.1101/2020.10.14.20212472. Accessed 2020-11-23

[31] Gavi, the Vaccine Alliance: Gavi COVID-19 situation report 12. https://www.gavi.org/sites/default/files/covid/Gavi-COVID-19-Situation-Report-12-20200630.pdf

[32] Gavi, the Vaccine Alliance: Gavi COVID-19 situation report 16. https://www.gavi.org/sites/default/files/covid/Gavi-COVID-19-Situation-Report-16-20200825.pdf

[33] Procter, S.R., Abbas, K., Flasche, S., Griffiths, U., Hagedorn, B., O’Reilly, K.M., Group, C.C.-.W., Jit, M.: SARS*CoV-2 infection risk during delivery of childhood vaccination campaigns: a modelling study. BMC Medicine 19(1), 198 (2021). doi:https://doi.org/10.1186/s12916-021-02072-8. Accessed 2021-10-21

[34] World Health Organization: Leave no one behind: guidance for planning and implementing catch-up vaccination (2021). https://www.who.int/publications/i/item/leave-no-one-behind-guidance-for-planning-and-implementing-catch-up-vaccination

[35] World Health Organization: Weekly epidemiological record (2021). http://apps.who.int/iris/bitstream/handle/10665/348056/WER9644-eng-fre.pdf

[36] World Bank: The COVID-19 Pandemic: Shocks to Education and Policy Responses. World Bank, Washington, DC (2020). doi:https://doi.org/10.1596/33696

[37] Taha, M.-K., Deghmane, A.-E.: Impact of covid-19 pandemic and the lockdown on invasive meningococcal disease. BMC Research Notes 13(1), 399 (2020). doi:https://doi.org/10.1186/s13104-020-05241-9

